# Antiviral innate immunity is diminished in the upper respiratory tract of severe COVID-19 patients

**DOI:** 10.1101/2022.11.08.22281846

**Authors:** Marcos J. Ramos-Benitez, Jeffrey R. Strich, Sara Alehashemi, Sydney Stein, Andre Rastegar, Adriana Almeida de Jesus, Farzana Bhuyan, Sabrina Ramelli, Ashley Babyak, Luis Perez-Valencia, Kevin M. Vannella, Gabrielle Grubbs, Surender Khurana, Robin Gross, Kyra Hadley, Janie Liang, Steven Mazur, Elena Postnikova, Seth Warner, Michael R. Holbrook, Lindsay M. Busch, Blake Warner, Willard Applefeld, Sarah Warner, Sameer S Kadri, Richard T Davey, Raphaela Goldbach-Mansky, Daniel S. Chertow

## Abstract

Understanding early innate immune responses to coronavirus disease 2019 (COVID–19) is crucial to developing targeted therapies to mitigate disease severity. Severe acute respiratory syndrome coronavirus (SARS-CoV)-2 infection elicits interferon expression leading to transcription of IFN-stimulated genes (ISGs) to control viral replication and spread. SARS-CoV-2 infection also elicits NF-κB signaling which regulates inflammatory cytokine expression contributing to viral control and likely disease severity. Few studies have simultaneously characterized these two components of innate immunity to COVID-19. We designed a study to characterize the expression of interferon alpha-2 (*IFNA2*) and interferon beta-1 (*IFNB1*), both type-1 interferons (IFN-1), interferongamma (*IFNG*), a type-2 interferon (IFN-2), ISGs, and NF-κB response genes in the upper respiratory tract (URT) of patients with mild (outpatient) versus severe (hospitalized) COVID-19. Further, we characterized the weekly dynamics of these responses in the upper and lower respiratory tracts (LRTs) and blood of severe patients to evaluate for compartmental differences. We observed significantly increased ISG and NF-κB responses in the URT of mild compared with severe patients early during illness. This pattern was associated with increased *IFNA2* and *IFNG* expression in the URT of mild patients, a trend toward increased *IFNB1*-expression and significantly increased *STING/IRF3/*cGAS expression in the URT of severe patients. Our by-week across-compartment analysis in severe patients revealed significantly higher ISG responses in the blood compared with the URT and LRT of these patients during the first week of illness, despite significantly lower expression of *IFNA2, IFNB1*, and *IFNG* in blood. NF-κB responses, however, were significantly elevated in the LRT compared with the URT and blood of severe patients during peak illness (week 2). Our data support that severe COVID-19 is associated with impaired interferon signaling in the URT during early illness and robust pro-inflammatory responses in the LRT during peak illness.

## Introduction

Pathogenesis of severe COVID-19 is driven by the interaction of SARS-CoV-2 infection and dissemination and the body’s attempt to contain and clear virus, resulting in direct and indirect cellular injury and dysfunction. SARS-CoV-2 infects the upper respiratory tract (URT) and lower respiratory tract (LRT) and replicates to high levels in type-2 pneumocytes lining alveoli (1). Early host cellular responses to SARS-CoV-2 infection include production of type 1 interferons (IFN-1), including interferon alpha-2 (*IFNA2*) and interferon beta-1 (*IFNB1*), and type 2 interferons (*IFNG*), after viral components including nucleic acids and proteins, termed pathogen-associated molecular patterns (PAMPs), are detected by pathogen recognition receptors (PRRs) on or within host cells (2). Interferons act in an autocrine and paracrine fashion to induce the expression of interferon-stimulated genes (ISGs), resulting in a local antiviral state (3-6). Beyond IFN responses, SARS-CoV-2 infection also induces pro-inflammatory responses, in part mediated by NF-κB (7), that regulate the expression of proinflammatory cytokines and chemokines and the activation and differentiation of innate and adaptive immune cells. While beneficial for viral control, NF-κB signaling might also contribute to immune-mediated organ injury and dysfunction (8).

Previous work suggests that impaired IFN responses are associated with severe COVID-19 and current knowledge in the field suggests innate and adaptive immune responses may vary across anatomical compartments and severity, perhaps related to cell types and microenvironments (9, 10). Furthermore, elevated cytokine and chemokine levels are thought to play a central role in severity and lethality in COVID-19 (11, 12). Thus, we aimed to characterize ISG and NF-κB response genes, central contributors to innate immunity in COVID-19, by illness severity and across anatomic compartments. We simultaneously assessed the trajectory of *IFNA2, IFNB1, IFNG*, ISGs, and NF-κB responses, using Nanostring technology (13), and SARS-CoV-2 RNA levels, by droplet digital polymerase chain reaction (ddPCR) assay, in the URT of patients with mild (outpatient) and severe (hospitalized) COVID-19. Among patients with severe COVID-19, we evaluated these parameters across the URT, LRT, and blood.

Our findings support early blunted IFN-1 mediated antiviral immunity in the URT of patients with severe COVID-19 and a robust NF-κB response in the LRT of these patients during peak illness. These observations are consistent with prior studies suggesting early impaired IFN signaling and robust pro-inflammatory responses in the LRT of patients with severe COVID-19.

## Methods

### Patient enrollment, clinical data, and samples collection

Patients with severe COVID-19 were admitted to the National Institutes of Health Clinical Center (NIHCC) and enrolled in the Adaptive COVID-19 Treatment Trial (ACTT-1) (#20-0006) between March 24-May 7, 2020, comparing remdesivir to placebo for the treatment of hospitalized patients with COVID-19 and evidence of lower respiratory tract disease. These patients were co-enrolled in the Admission and Management of Occupational or Other Exposures to Biodefense/Bioterrorism Agents or to Epidemic/Emerging Infectious Diseases (AMOEBAE) protocol (#10-I-0197), allowing for serial collection of blood and respiratory specimens. Samples were collected near daily from the URT and blood of all patients with severe COVID-19 and intermittently from the LRT of mechanically ventilated patients. Clinical data were captured in a REDCAP database, including demographics, comorbidities, laboratory values, supportive care, and outcomes. Outpatients with mild COVID-19 were enrolled in the National Institute of Health Transmissibility and Viral Load of SARS-CoV-2 Through Oral Secretions Study (Study Number, NCT04348240, NIH IRB 20-D-0094). Healthy control blood samples were collected under the National Institute of Health approved protocol Number: 17-I-0016 (NCT02974595): Studies of the Natural History, Pathogenesis, and Outcome of Autoinflammatory Diseases (NOMID/CAPS, DIRA, CANDLE, SAVI, NLRC4-MAS, Still-like Diseases, and other Undifferentiated Autoinflammatory Diseases). Deidentified healthy control respiratory samples were provided by the Department of Laboratory Medicine at the NIHCC. These samples that were fully deidentified to all study team members were deemed non-human subject research by the NIH Office of Human Subject Research Protection (OHSRP) in accordance with Federal Policy for the Protection of Human Subjects at 45 CFR 46. Sample collection and analyses from patients with mild and severe COVID-19 are represented in **Figure 1**. All patients ID used in the manuscript are not identifiable by anyone outside the research group.

**Figure 1.**
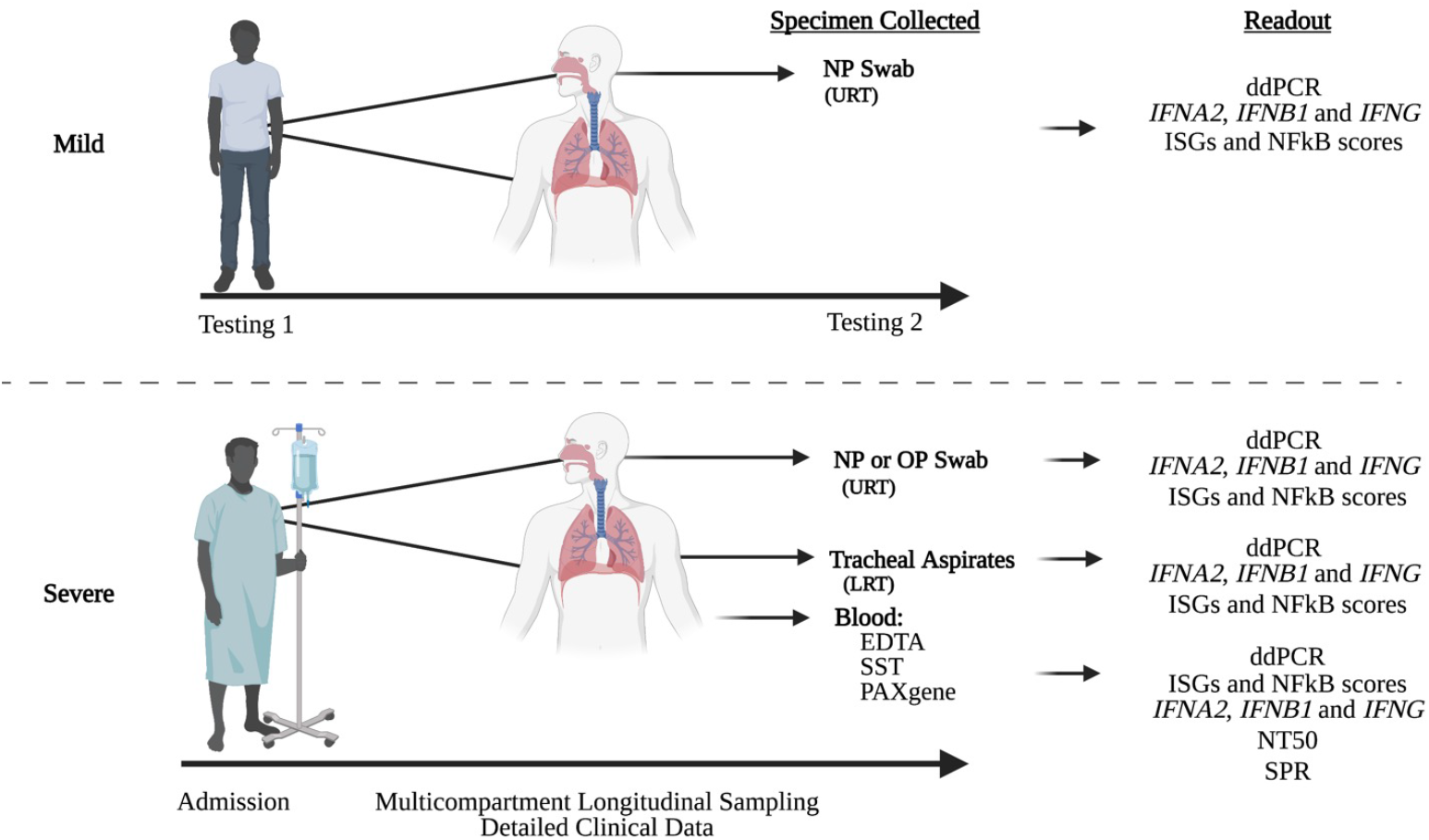
Study design diagram.

### Quantification of SARS-COV-2 RNA in respiratory samples by ddPCR

Total RNA was extracted from oropharyngeal and nasopharyngeal swabs preserved in viral transport media and plasma using the QIAamp Viral RNA Mini Kit (Qiagen, Germantown, MD, USA) according to the manufacturer’s protocols. Total RNA was extracted from tracheal aspirates digested 1:1 with Remel Sputasol (ThermoFisher Scientific, Waltham, MA) according to manufacturer’s instructions, and 200 μL of digested tracheal aspirate were added to 1 mL QIAzol (Qiagen). A chloroform extraction was performed, and the aqueous layer was added to the RNeasy Mini Kit (Qiagen) to extract total RNA using the manufacturer’s protocol for animal cells and tissues. The NanoDrop ND-1000 Spectrophotometer (Thermo Scientific, Waltham, MA, USA) was used to quantify RNA concentrations from tracheal aspirates. RNA extracted via the QIAamp Viral RNA Mini Kit was not quantified via NanoDrop due to the presence of carrier RNA. The QX200 AutoDG Droplet Digital PCR System (Bio-Rad, Hercules, CA, USA) was used to detect and quantify SARS-CoV-2 RNA using the SARS-CoV-2 ddPCR Kit (Bio-Rad) as previously described (14)

### Determining ISGs and NF-κB gene responses in respiratory samples and blood by NanoString assay

Total RNA was extracted from oropharyngeal, nasopharyngeal swabs and/or tracheal aspirates, as described above. Total RNA was extracted from whole blood samples collected in PaxGene tubes (Qiagen, Germantown, MD). Gene expression of selected genes was determined by NanoString assay (NanoString Technologies, Seattle, WA) and a 28-gene type I ISGs score (13, 15) were calculated as described. Briefly, the 28-gene type I ISGs score is the geometric mean expression of 28 IFN-stimulated genes (ISGs), and the 11-gene NF-κB score is the geometric mean expression of 11 NF-κB targets. ISGs and NF-κB genes included in the respective scores are listed in **Figure S1**.

### Characterization of antibody responses by surface plasmon resonance

The SARS-CoV-2 Spike plasmid expressing genetically stabilized pre-fusion 2019– nCoV_S–2P spike ectodomain, a gene encoding residue 1-1208 of 2019–nCoV S fused to 8xHisTag, was a kind gift from Barney Graham (Vaccine Research Center, NIH). This expression vector was used to transiently transfect FreeStyle293F cells (ThermoFisher) using polyethyleneimine. Protein was purified from filtered cell supernatants using StrepTactin resin and subjected to additional purification by size–exclusion chromatography in PBS. Steady-state equilibrium binding of post-SARS-CoV-2 infected human polyclonal plasma was monitored at 25°C using a ProteOn surface plasmon resonance (BioRad). The purified recombinant SARS-CoV-2 prefusion spike protein was captured via a His-tag to a Ni-NTA sensor chip with 200 resonance units (RU) in the test flow channels. The protein density on the chip was optimized such as to measure monovalent interactions independent of the antibody isotype (12, 16, 17). Serial dilutions (10-, 50- and 250-fold) of freshly prepared plasma in BSA-PBST buffer (PBS pH 7.4 buffer with Tween-20 and BSA were injected at a flow rate of 50 μL/min (120-sec contact duration) for the association, and disassociation was performed over a 600-second interval. Responses from the protein surface were corrected for the response from a mock surface and for responses from a buffer-only injection. Surface plasmon resonance (SPR) was performed with the serially diluted plasma of each individual time point in this study.

### Plaque reduction neutralizing assays

Serum samples were tested for neutralizing antibodies using a fluorescence reduction neutralization assay (FRNA). Briefly, the test material was diluted in the serum-free medium through a 12-step 2-fold serial dilution (1:20-1:81920) and then mixed with approximately 4.54 log_10_ of SARS-CoV-2 (2019-nCoV/USA-WA1-A12/2020 from the US Centers for Disease Control and Prevention, Atlanta, GA, USA) to achieve a multiplicity of infection of 0.5 in 96-well plates. Following a 1h incubation at 37ºC/5% CO_2_, the virus-serum mixture was added to washed Vero E6 cells (BEI) and incubated for 24h at 37ºC/5% CO_2_ to allow virus infection. Following incubation, the plates were fixed overnight at 4ºC with neutral-buffered formalin and removed from the biocontainment laboratory. The cells were washed with PBS and permeabilized with 0.25% Triton-X100 in PBS for 10 min at room temperature. The cells were washed three times with PBS and then incubated in a blocking buffer (3% BSA in PBS) for 30 min at room temperature. The cells were washed three times with PBS, and an anti-SARS-CoV-1 nucleoprotein primary rabbit antibody (Sino Biological) diluted in 3%BSA/PBS was added to the plates and incubated for 1h at room temperature. The plates were washed three times with PBS, followed by adding an AlexaFluor conjugated goat anti-rabbit secondary antibody (Life Technologies) diluted in PBS containing the Hoechst 33342 nuclear stain. Cells were incubated for 30 minutes at room temperature in the dark, washed with PBS, and then PBS was added back to the plates to prevent drying. Individual infected cells were counted using an Operetta high content imaging system (Perkin-Elmer). Four fields of view were counted in each well with a minimum of 1000 cells per field. Each sample was tested in duplicate-on-duplicate plates for a total of ∼16000 cells counted per dilution for each test sample. A four-parameter logistical regression was performed on individual dilution series (Prism 8.0, GraphPad) to determine the IC_50_ for each serum sample

### Statistical analysis

We used the Kruskal-Wallis test or Unpaired T test for group comparisons, p-values below 0.05 as statistically significant. Statistical analyses and graphing were done using R-studio version 1.3 and GraphPad Prism 9.

## Results

### Severe and mild cohort description

Among eight severe patients included in this study, the mean age was 48.1 years (range 22-67), 75% were male, 50% were White, 25% were Black/African American, and 25% were Asian. The mean maximum Sequential Organ Failure Assessment (SOFA) score was 5 (range 1 to 12). Four patients required invasive mechanical ventilation, three required high flow nasal cannula, and one required low flow oxygen by nasal canula during their hospitalization (**Table S1)**. The mean length of hospital stay was 18 days (range 6 to 27). Seven of the eight severe patients survived through hospital discharge. A summary of COVID-19 specific therapies is provided in **Table S2**. Among the seven mild patients, the mean age was 31.7 years (range 21-59), 14% were male, 86% were White, and 14% were Asian **(Table S3)**.

### Higher ISGs and NF-kB genes scores in the URT of mild compared with severe COVID-19 patients during early illness

URT samples were available from six of seven mild patients (M1-M5 and M7) and four of eight severe patients (S2-S5) within 10 days (D) of illness onset (early illness) for comparison **(Tables S1 and S3)**. For this analysis, the average day of URT sample collection following illness onset in the mild and severe cohort was D5 and D8, respectively. The mean URT ISGs and NF-κB genes scores were significantly elevated (P<0.05) in mild compared with severe COVID-19 patients and healthy controls during early illness (**Figure 2)**. No difference in URT ISGs and NF-κB scores were observed in severe COVID-19 patients relative to healthy controls.

**Figure 2.**
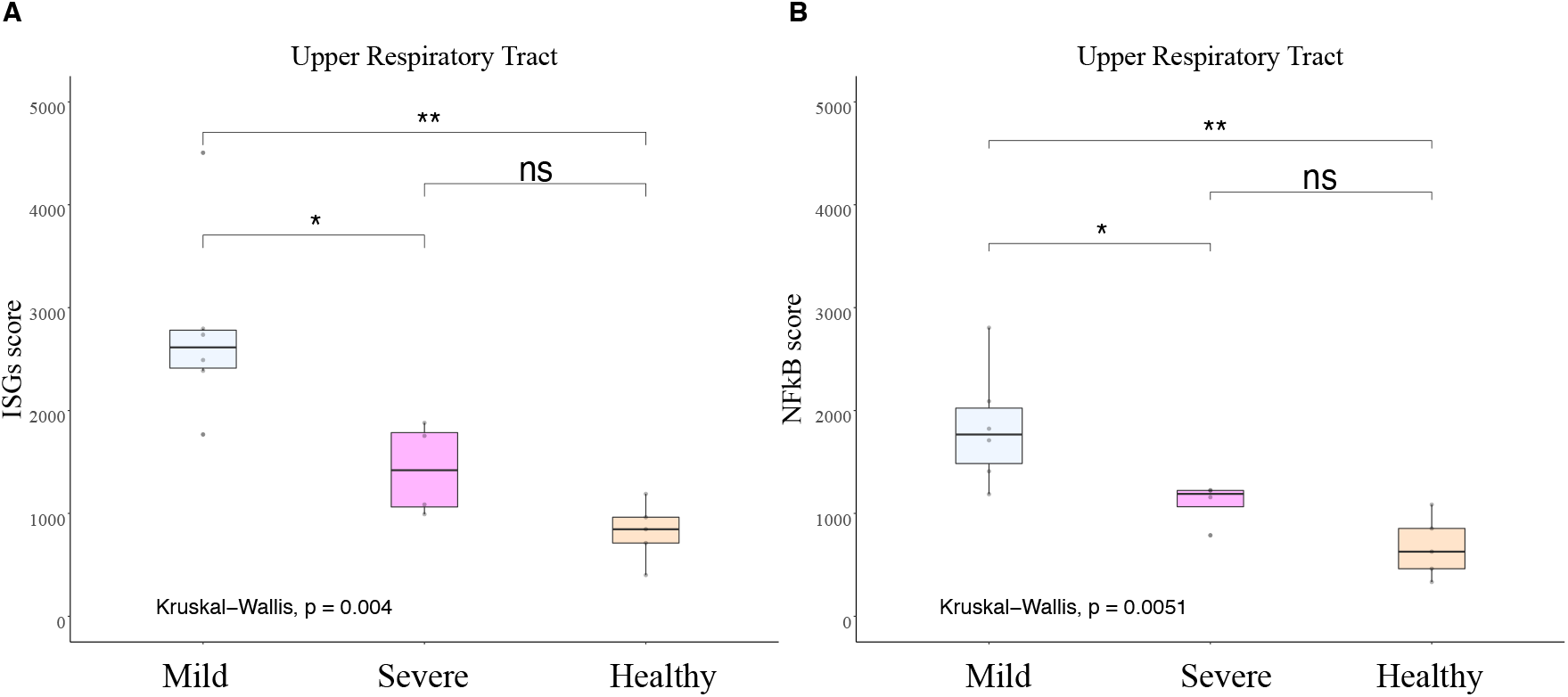
Higher ISGs and NF-κB in the URT in mild COVID-19 patients. ISG Score and NF-kB score measured by NanoString in upper respiratory samples collected in the first 10 days from the post-symptom onset in mild (n=6) and severe COVID patients (n=4) compared to healthy control (n=5). **A**) ISGs score measured by NanoString in upper respiratory samples collected in the first 10 days from the post-symptom onset. **B**) NF-κB score measured by NanoString in upper respiratory samples collected in the first 10 days from the post-symptom onset. For both scores, mild and severe COVID-19 patients were compared to healthy control (n=5). Kruskal Wallis test is used to compare the three groups and corrected Dunn’s test for multiple comparisons was used to compare mild and severe COVID patients to HC. \**P*<0.05, \*\**P*<0.01, NS *P*<0.05

To better understand individual gene contributions to ISGs and NF-κB genes scores, we compared expression levels of individual gene components of these scores between mild and severe COVID-19 patients normalized to expression levels in healthy controls. Significantly higher expression of 13 type I ISGs, was observed in the URT of mild compared with severe COVID-19 patients during early illness, including *CXCL10, DDX60, GBP1, HERC5, HERC6, IFIT5, OAS3, OASL, RTP4, SIGLEC1, SOCS1, SPATS2L, USP18* (P<0.05) **(Figure 3)**. Analysis of individual gene components of the NF-κB score revealed that expression levels of *AICDA, IFNG, TANK*, and *TLR2* were significantly increased in the URT of mild compared with severe COVID-19 patients during early illness, while only *EBI3* had significantly higher expression in severe COVID-19 patients (**Figure S2)**.

**Figure 3.**
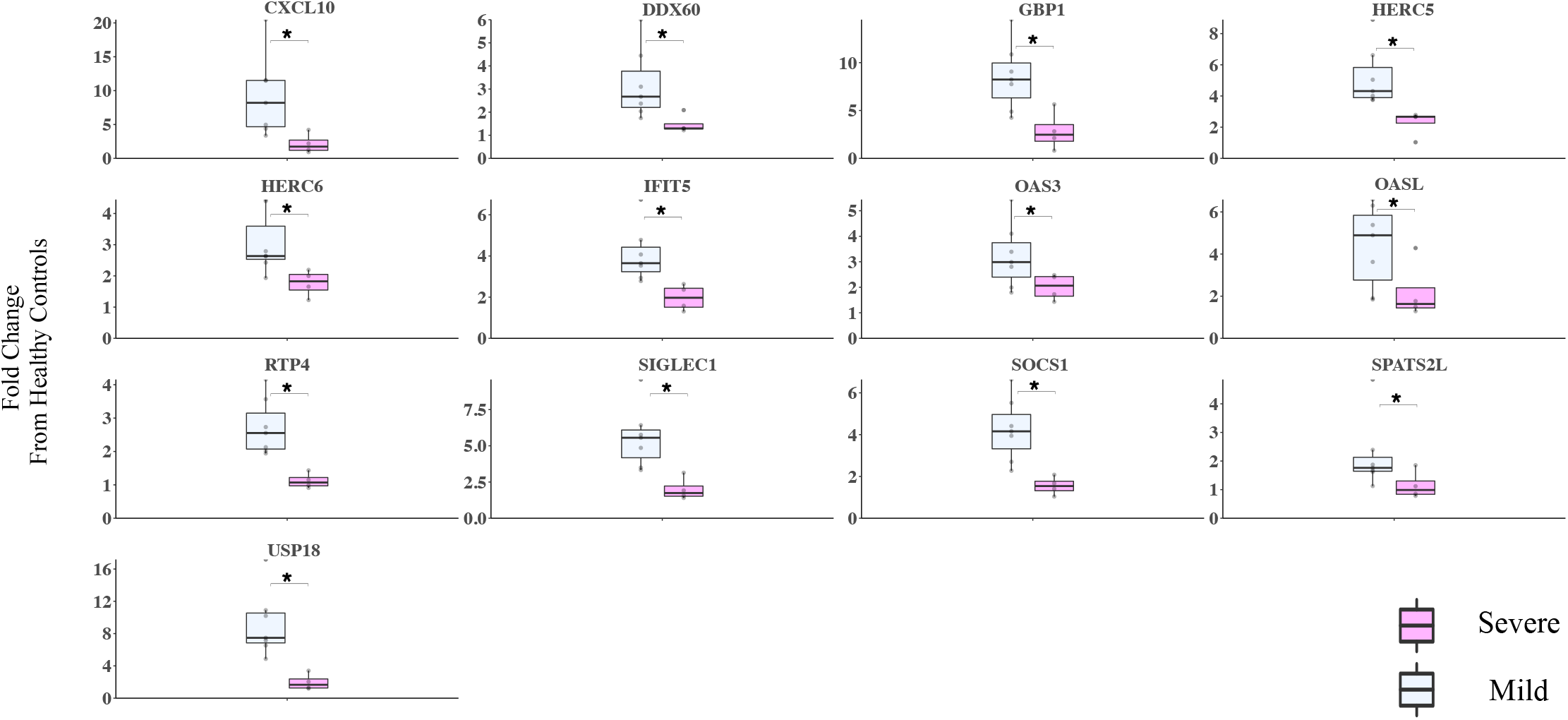
Thirteen type I IFN response genes are increased in mild COVID-19 in the URT. The 13 type I IFN response gene transcriptions were measured by NanoString in the upper respiratory tract of patients in the first 10 days from the post-symptom onset. mild (n=6) and severe COVID (n=4) patients. Data for both groups are shown as fold change from healthy controls. Kruskal Wallis test is used to compare mild and severe COVID patients, \**p*<0.05,

### Quantification of SARS-CoV-2 RNA in respiratory samples of mild and severe COVID-19 patients and antibody responses in blood of severe COVID-19 patients

We measured SARS-CoV-2 RNA levels in the URT from five mild (M1-M5) and four severe COVID-19 patients (S2-5) who had samples collected within and after D10. Individual patient data are presented in **Table S4**. We observed limited control of viral load in the URT of mild and severe COVID-19 patients **(Figure 4)** and rising viral RNA levels in the LRT of severe patients after D10 (**Figure S3**). LRT samples from mild patients were not available for analysis. Anti-spike protein antibodies titers (MaxRU) and neutralizing capacity (NT_50_) peaked in plasma of severe COVID-19 patients between D14 and D21 and subsequently plateaued **(Figure S4). Higher IFNA2 and IFNG expression, but not IFNB1 expression, in the URT of mild compared with severe COVID-19 patients during early illness**

**Figure 4.**
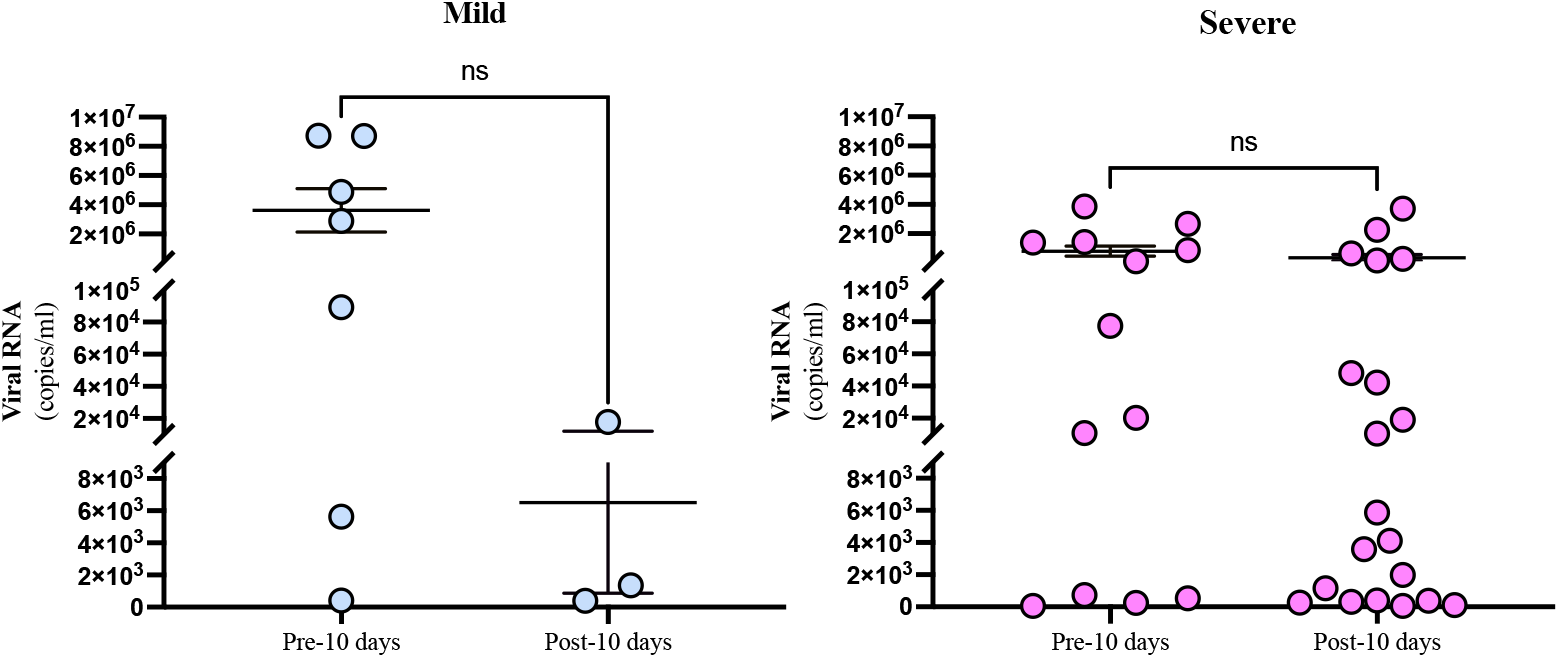
Viral load in the URT of mild and severe patients. Viral load was measured by ddPCR from patients that had samples before and after 10 days post-symptoms onset. **A**) Shows viral load in the mild subjects first 10 days post-symptom onset (n=5, 7 samples) compared with after day 10 post-symptom onset (n=3, 3 samples), p=0.1629. **B**) Shows viral load in the severe subjects for the first 10 days post-symptom onset (n=4, 13 samples) compared with after day 10 post-symptom onset (n=4, 20 samples), p=0.2527. Individual dots indicate independent samples, solid line indicates the mean and SEM. Unpaired T test was used to compared between groups.

We hypothesized that differences in the upstream expression of *IFNA2, IFNB1* and *INFG* between mild and severe COVID-19 patients might account for significant differences in downstream ISGs scores during early illness. To test this, we compared URT gene expression of these targets in mild and severe COVID-19 patients expressed as a fold-change from healthy controls. We observed significantly higher expression of *IFNA2* and *IFNG* (p<0.05), but not *IFNB1* in the URT of mild compared with severe COVID-19 patients during early illness **(Figure 5A-C)**. To better contextualize the opposite trend in *IFNB* expression we evaluated gene expression levels of cyclic GMP–AMP (cGAS) and cytosolic sensor stimulator of interferon genes (STING), which are upstream mediators of IFNβ expression through the cGAS–STING signaling axis (18). Both targets were significantly higher in severe than mild COVID-19 patients (P<0.05) (**Figure 5D-E)**.

**Figure 5.**
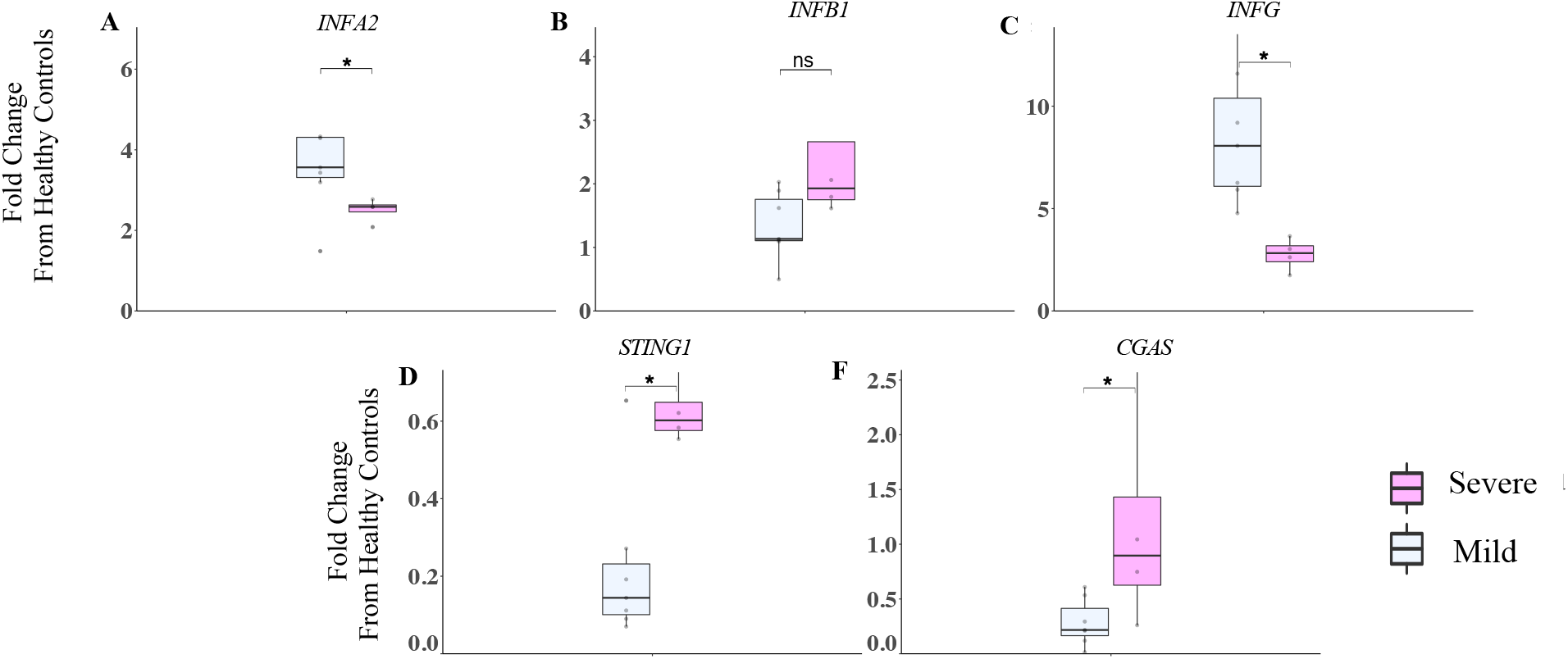
Increased expression of CGAS/STING INFB1 pathway in the URT of severe COVID-19 patients. A – F)The IFNA2, IFNB1, IFNG, STING-1, and CGAS gene transcriptions were measured by NanoString in the URT of patients in the first 10 days following symptom onset. Mild (n=6) and severe COVID (n=4) patients. Data for both groups are shown as fold change from healthy controls. Kruskal Wallis test is used to compare mild and severe COVID patients \**P*<0.05, NS *P*>0.05

Interferon regulatory factors (IRFs) are a family of transcription factors that regulate IFN1 gene induction downstream of PRRs and drive anti-viral and pro-inflammatory responses (19). We evaluated a subset of IRFs (IRF3, IFR5, and IRF7-IRF9) in mild versus severe Covid-19 patients to characterize differences in URT gene expression. We observed that IRF3 was significantly higher in severe compared to mild COVID-19 patients (**Figure 6A**) (p<0.05). Conversely, IRF5 and IRF8 were significantly higher in mild than severe COVID-19 patients (**Figure 6B and D**) (*P*<0.05).

**Figure 6.**
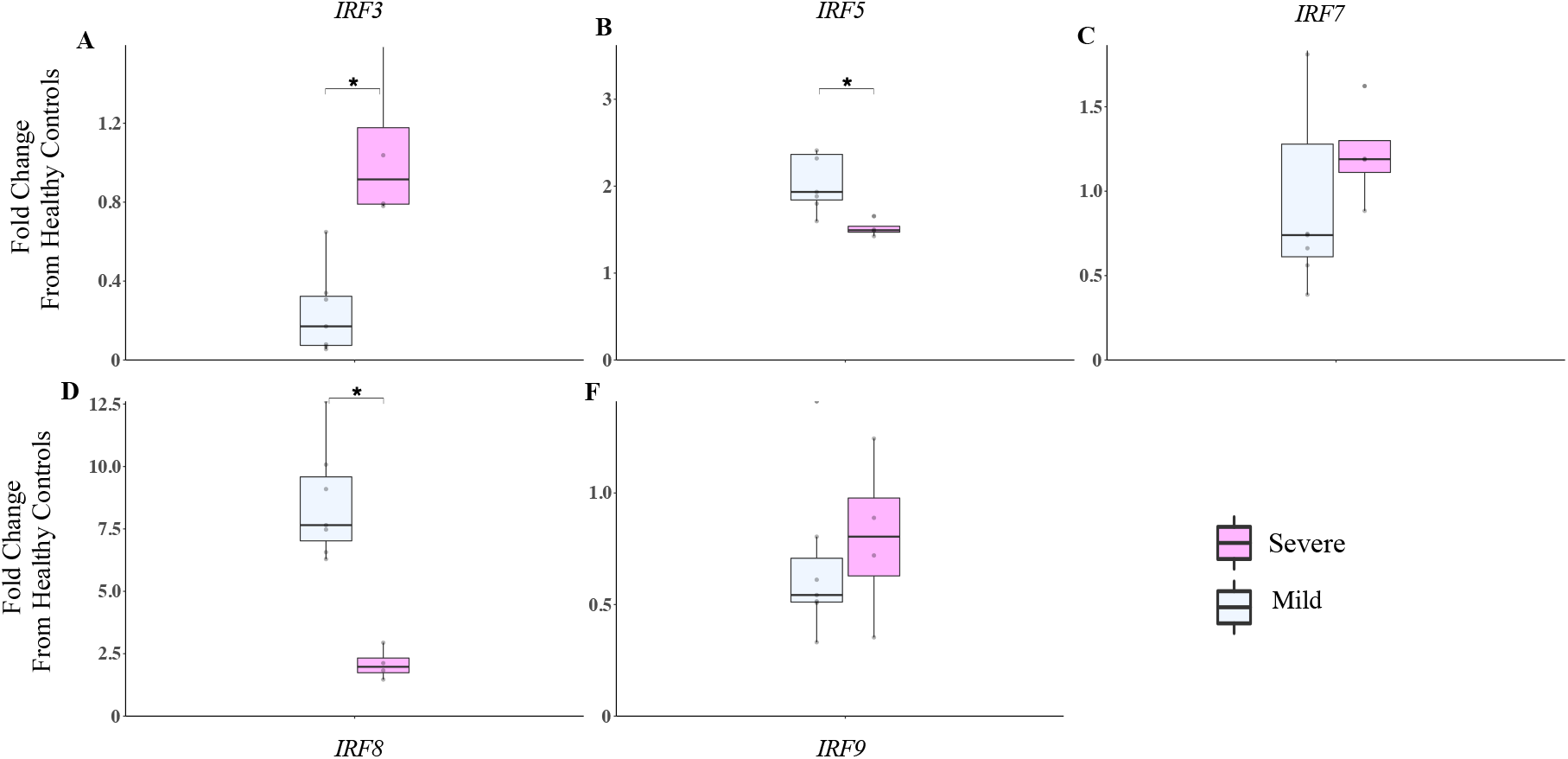
IRFs are differentially regulated in mild and severe COVID-19 patients. **A-F**) The IRF3, IRF5, IRF7, IRF8 and IRF9 gene transcriptions were measured by NanoString in the URT of patients in the first ten days following symptom onset. Mild (n=6) and Severe COVID (n=4) patients. Data for both groups are shown as fold change from healthy controls. Kruskal Wallis test is used to compare Mild and severe COVID patients \**P*<0.05

### Across anatomic compartment and by week comparison of IFN, ISGs, and NF-κB gene expression among severe COVID-19 patients

Among patients with severe COVID-19 we collected time-matched samples from the URT, LRT, and blood and characterized differences in IFN, ISGs, and NF-κB genes expression by illness week. We observed higher expression of *IFNA2, IFNB*1and *IFNG* in the respiratory tract compared to blood during illness weeks one through three (p<0.05) **(Figure 7 A – C)**. We also observed significantly higher expression of *IFNB*1 and *IFNG* in the URT versus LRT during week two. During week 1, we observed significantly higher ISGs scores in blood than URT. ISG expression in blood significantly decreased over time **(Figure 7D)**. We observed significantly higher NF-κB scores in the LRT relative to the URT and the blood during week two (*P*<0.01, *P*<0.0001, respectively) and in the URT relative to the blood in week two (P<0.05) (**Figure 7E)**.

**Figure 7.**
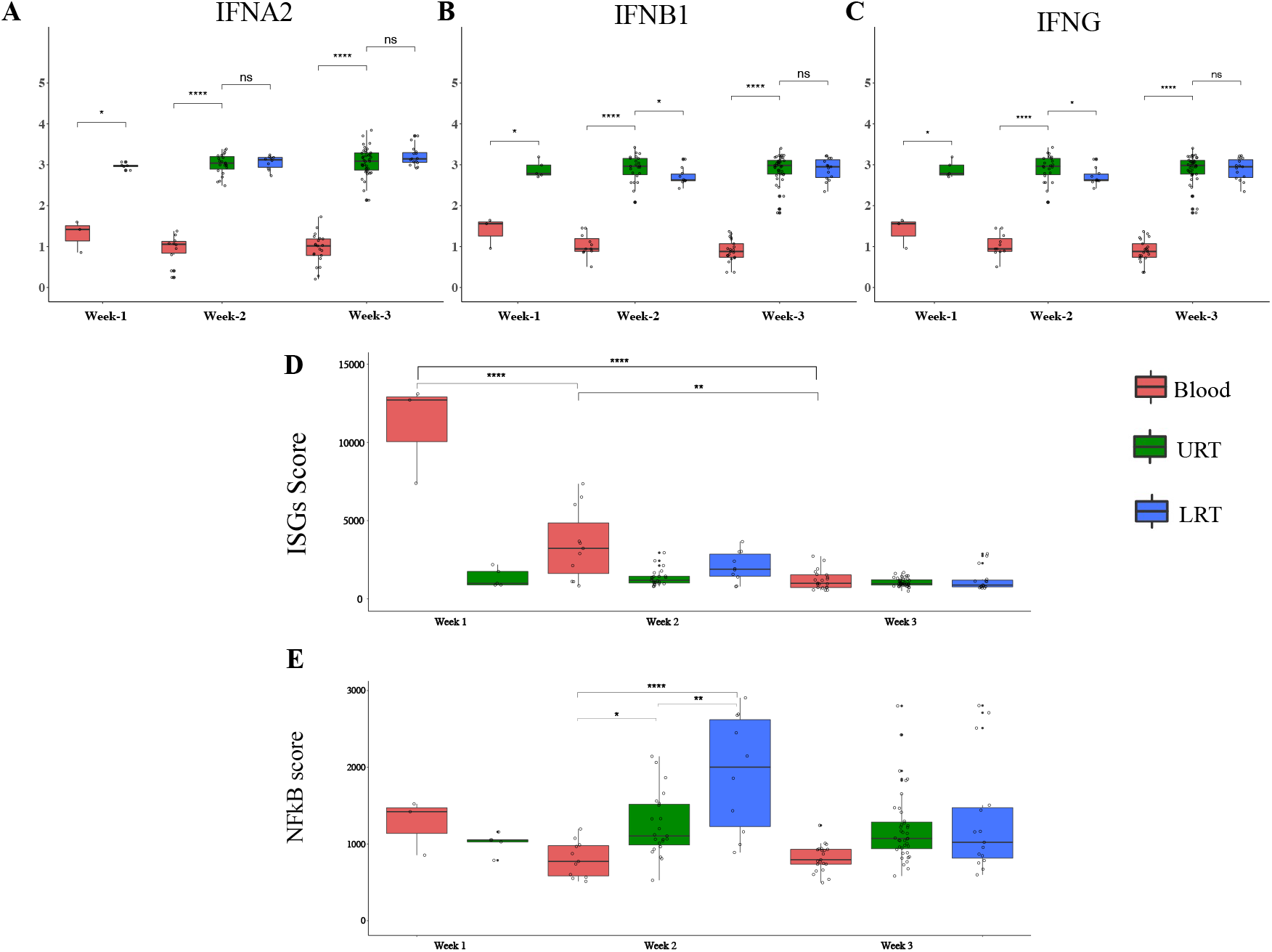
ISGs and NF-κB scores differ across anatomical compartment in severe COVID-19 patients. **A-C**) *IFNA2, INFB1* and *IFNG* expression measured by NanoString in upper respiratory samples collected in the first three weeks following symptom onset in severe COVID-19 patients compared to healthy control (n=5). **D-E**) ISGs and NF-κB score were measured by NanoString in Blood (Week 1 n=3, Week 2 n=11, Week3 n=22), URT (Week 1 n=5, Week 2 n=23, Week3 n=39), and LRT (Week 2 n=10, Week3 n=15). Kruskal Wallis test is used to compare the three groups and corrected Dunn’s test for multiple comparisons was used to compare mild and severe COVID patients to HC. \**P*<0.05, \*\**P*<0.01, \*\*\**P*<0.001, \*\*\*\**P*<0.0001

## Discussion

In this work, we assessed SARS-CoV-2 RNA levels, IFN-1, ISGs, and NF-κB genes responses within the respiratory tract of mild and severe COVID-19 patients and blood of severe COVID-19 patients to inform host-pathogen interactions among these cohorts during illness. Lower ISGs and NF-κB genes scores in the URT of severe compared with mild COVID-19 patients suggests impaired early innate immune responses in the URT of severe COVID-19 patients. Viral proteins or host factors can disrupt intercellular mediators of innate immune responses. For example, blocking signal transducer and activator of transcription (STAT) phosphorylation or nuclear translocation has been implicated in impaired IFN-I responses in severe COVID-19 (20-22).

Among the ISGs we studied, 13 genes were increased in the URT in mild compared to severe COVID-19 patients. Most of these 13 genes have an antiviral or regulatory role, summarized in **Table S5**. A clear example is CXCL10, where we observed the greatest difference in the URT of mild versus severe COVID-19 patients. The *CXCL10*/*CXCR3* axis plays a non-redundant role in coordinating early adaptive immune responses through recruitment of CXCR3+ effector T and NK cells (23, 24). Decreased expression of *CXCL10* in the URT of severe patients might limit NK and T cell recruitment, impairing recognition and clearance of SARS-CoV-2 infected cells (25). Our observations are consistent with previous single-cell RNA sequencing analysis of URT samples from COVID-19 patients showing adequate IFN-1 and anti-viral gene responses among mild/moderate COVID-19 patients and recruitment of myeloid but not T cells in severe patients (26).

To better understand the difference in ISGs response, we evaluated expression levels of *IFNA2, IFNB1*, and *INFG*. We observed that *IFNA2* and *IFNG* were significantly elevated in the URT of mild compared with severe COVID-19 patients, suggesting an intact and appropriate early antiviral response in the mild but not severe patients. Robust innate immune responses in the URT of the mild COVID-19 patients may have limited spread or burden of SARS-CoV-2 to the LRT of these patients, although we were unable to directly assess this given LRT samples were not available from mild COVID-19 patients. In contrast, impaired responses in the URT of the severe COVID-19 patients were associated with increasing SARS-CoV-2 burden in the LRT after day 10 of illness. This finding is consistent with prior studies showing more rapid clearance of SARS-CoV-2 among mild compared with severe COVID-19 patients (25, 27). *IFNB1* gene expression trended higher in severe compared with mild COVID-19 patients. Consistent with this, we observed significantly higher expression of cGAS-STING pathway genes in severe compared with mild COVID-19 patients. This increased *cGAS* and *STING* expression in severe COVID-19 patients suggests an upregulation of the cGAS-STING-IFNβ axis, which has recently been associated with endothelial dysfunction in severe COVID-19 (28).

We further explored the differences in *IFNA2, IFNB1*, and *INFG* responses in mild compared to severe COVID-19 patients by assessing differences in expression of upstream regulators, IFRs. *IRF3* expression was higher in severe compared with mild COVID-19 patients, which aligns with cGAS, STING and *IFNB1* being higher in severe COVID-19 patients. Studies have shown that overexpression of cGAS activates IRF3 and induces interferon-β expression through STING (29). Conversely, IRF5 was significantly higher in mild compared with severe COVID-19 patients. Previous studies have shown that an IRF5 deficiency impairs secretion of IL-1β, IL-6, IL-12, IL-23 and TNFα and skewing of T cell subsets from T helper 1 (Th1) and T helper 17 (Th17) responses (30, 31), that contribute to clearance of intracellular pathogens, toward T helper 2 (Th2) cells (30). In severe COVID-19, the predominance of Th2-associated cytokines, including IL-4, IL-5, IL-13, and IL-10 have been associated with delayed viral clearance (32). IRF5 signaling in our mild cohort is consistent with intact early Th1 response in the URT of these patients, perhaps limiting the progression of illness to the LRT. IRF8 was also higher in the mild than in the severe COVID-19 patients. IRF8 is expressed in Treg and Th1 cells and is associated with CXCR3 expression (33). IRF8 also controls the survival and function of terminally differentiated monocytes, Type-I conventional dendritic cells, and plasmacytoid dendritic cells. Conversely, IRF8 suppresses neutrophil generation (34). Consistent with this finding, increased absolute neutrophil counts have been associated with severe but not mild COVID-19 (35). These data are consistent with the notion that in mild patients, viral infection may directly stimulate appropriate selected ISGs responses, including *CXCL10*, and activate IFNa production that bridges innate and adaptive immunity through skewing DC differentiation/activation, towards the priming and expansion of Th1 cells, i.e. in the nasal- or nasopharynx-associated lymphoid tissue (NALT) in the URT, perhaps contributing to improved viral clearance while maintaining homeostasis and self-tolerance following a robust immune response (36).

NF-κB responses that were lower in the URT of severe compared with mild COVID-19 patients were increased in the LRT of severe patients at week 2. Exuberant NF-κB responses have been previously associated with and are thought to contribute to immunopathogenesis of lung injury in COVID-19 (37). Our findings are consistent with this, as week 2 represented peak illness among our severe COVID-19 patients. Peak humoral responses by week 3 in these patients coincided with resolution of exuberant NF-κB responses, consistent with effective adaptive immunity contributing to disease resolution.

To characterize innate immune responses in blood versus respiratory tract among severe patients, we evaluated *IFNA2, IFNB1, IFNG*, ISGs, and NF-κB genes responses across compartments by week of illness. We observed significantly elevated *IFNA2, IFNB1*, and *IFNG* expression in the respiratory tract compared with blood throughout illness. Despite higher IFN expression in the respiratory tract compared with blood, we observed significantly higher ISGs responses in the blood compared with the URT during week 1. Higher ISGs responses in blood during week 1 among severe COVID-19 patients is consistent with an early circulation of activated myeloid cells (monocytes and neutrophils) that are subsequently recruited to the lungs of severe patients. Lower ISGs responses in the URT compared with blood, despite higher IFN expression in the URT, is consistent with impaired IFN signaling in SARS-CoV-2 infected nasopharyngeal epithelium among severe patients through host and viral factors that directly suppress ISGs and the IFN response (38, 39). These findings are consistent with observations from prior studies (26).

Generalizability of our study findings are limited by our small sample size and the fact that all patients were infected prior to availability of vaccines and circulation of SARS-CoV-2 variants. Baseline differences in demographics, comorbid conditions, and the exact timing of sample collection between our mild and severe cohorts might contribute to observed differences in our study. However, differences in IFN and NF-κB responses between our cohorts provide insight into effective compartmental innate immune responses in the nasopharynx associated of younger patients, compared with less effective responses among older patients with pre-existing conditions. Differences in innate immune responses in the blood and respiratory tract of severe patients and evolution of these responses over time provide insight into the compartmental differences in the molecular pathogenesis of severe COVID-19 and support pursuit of similar larger studies to help guide development of improved prognostic markers and host-targeted therapies for severe COVID-19.

## Data Availability

Data produced in the present work are contained in the manuscript and supplementary figures.

## Author Contributions

D.S.C, R.GM., M.J.R.B conceived and supervised the study. J.R.S, G.G K.H, J.L, S.M, E.P, Sa.W, S.M, E.P, L.M.B managed and/or collected clinical data. S.S S.A,, A.R, A.dJ, F.B, S.R, A.B, L.P.V, K.M.V, G.G, S.K, R.B, S.W, M.R.H, L.M.B, B.W, W.A, S., S.K, R.T.D and R.G.M collected, processed samples and/or analyzed experimental data. M.J.R.B wrote the manuscript. All authors reviewed and edited the manuscript and approved the final version.

## Acknowledgments

We thank members of the Sjogren’s Clinical Investigations Team for collecting specimens and data.

## Notes

All claims expressed in this article are solely those of the authors and do not necessarily represent those of their affiliated organizations, or those of the publisher, the editors and the reviewers. Any product that may be evaluated in this article, or claim that may be made by its manufacturer, is not guaranteed, or endorsed by the publisher.

