## Supplementary Figures for "Antiviral innate immunity is diminished in the upper respiratory tract of severe COVID-19 patients"

### Supplementary Figure 1

| 28 Type I Interferon Stimulated Genes | |
|---|---|
| EPSTI1 | IFI44 |
| IFIT1 | IFI44L |
| IFIT2 | IFI6 |
| IFIT3 | IFIT5 |
| ISG15 | LAMP3 |
| LY6E | MX1 |
| USP18 | OAS1 |
| CXCL10 | OAS2 |
| SOCS1 | OAS3 |
| GBP1 | OASL |
| DDX60 | RSAD2 |
| HERC5 | RTP4 |
| HERC6 | SIGLEC1 |

| NF-kB enriched Score |
|---|
| AICDA |
| CCND2 |
| EBI3 |
| GZMB |
| IFNG |
| MSR1 |
| SELL |
| SELP |
| TANK |
| TLR2 |
| XIAP |

IFNα and IFNγ regulated genes

IFNα, IFNγ and NFκB1/2 regulated genes

IFNα and NFκB1/2 regulated genes

^Liu SY et al, PNAS, 2012

**Figure S1.** List of genes contained in the IFN I and NF-κB score. Calculation of an IFN score by 2 methods: a z-score-based standardized score and a geomean score. Calculation of an NF-κB score by 2 methods: a z-score-based standardized score and a geomean score

### Supplementary Figure 2

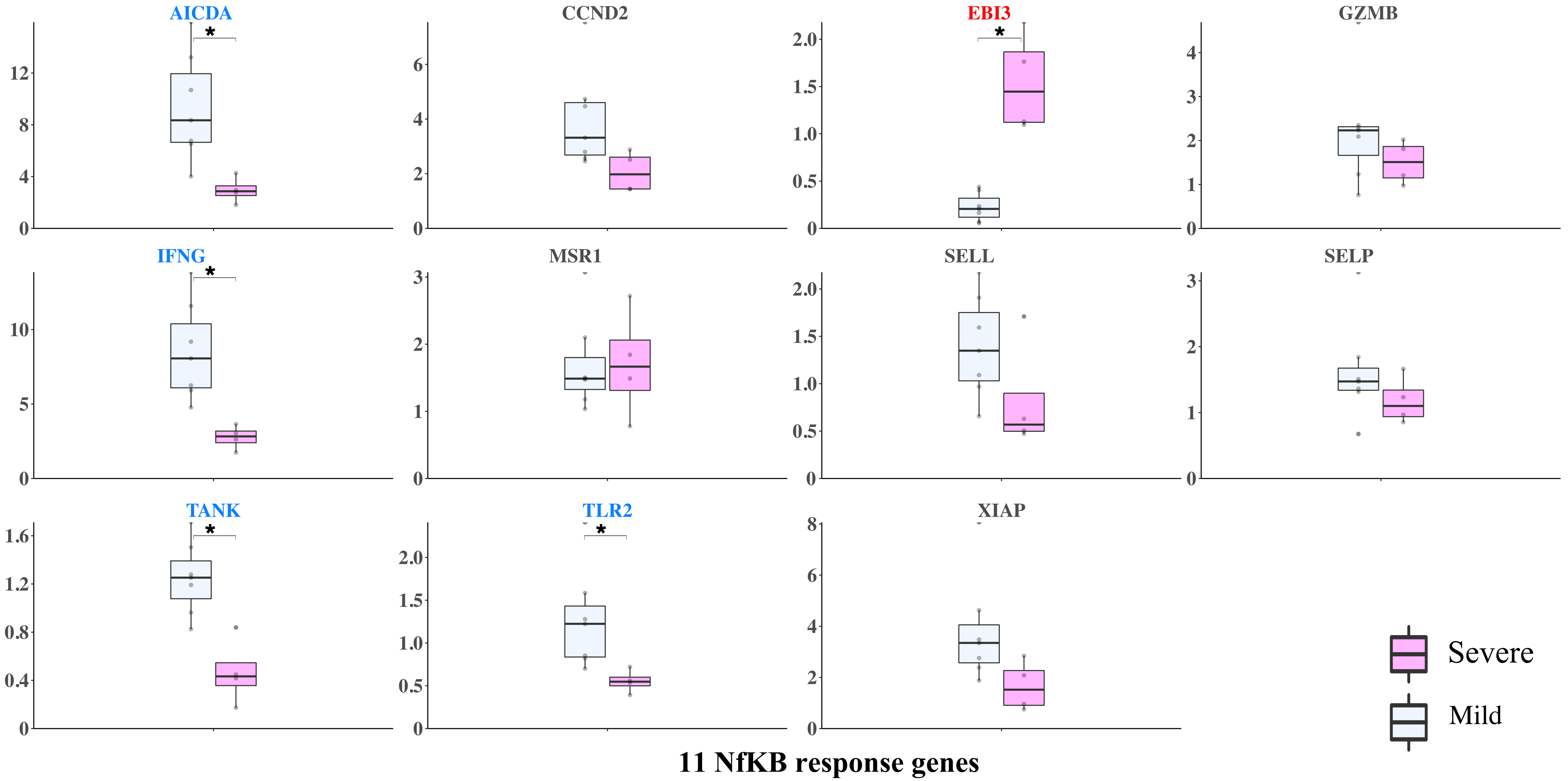


**Figure S2.** The 11 NF-κB response gene transcriptions were measured by NanoString in the upper respiratory tract (URT) of patients in the first ten days from the disease onset. mild (n=6 patients) and severe COVID (n=4 patients, the earliest sample is included). Data for both groups are shown as fold change from healthy controls. Blue labels indicate gene expression was significantly higher in the mild than in the severe cohort, Red label indicates gene expression was significantly higher in the severe than in mild cohort. *$P$<0.05, **$P$<0.01, ***$P$<0.001, ****$P$<0.0001.

### Supplementary Figure 3

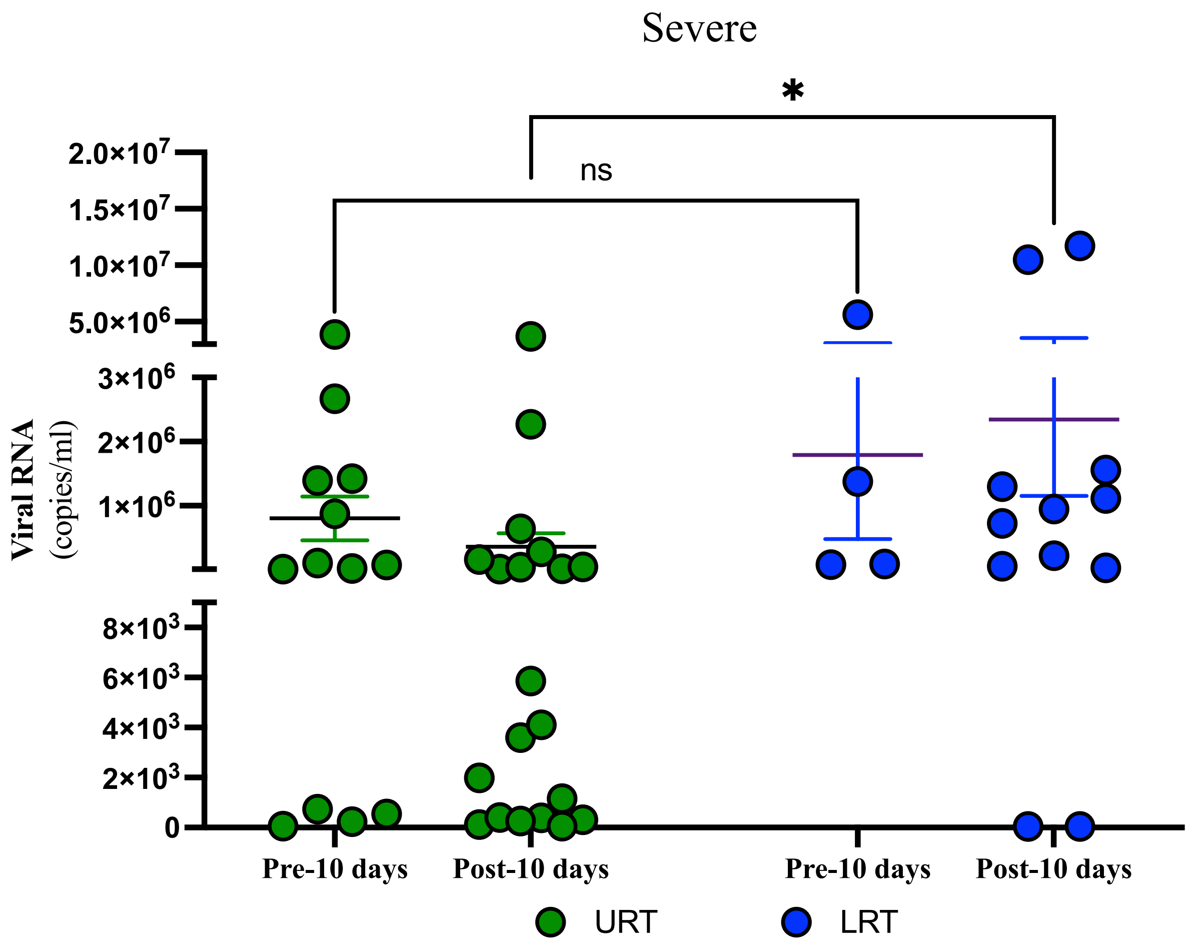


**Figure S3.** Viral load was measured by ddPCR from severe COVID-19 patients that had samples before and after 10 days post-symptoms onset. In the URT for the first 10 days, (n=4, 13 samples), post 10 days (n=4, 20 samples). In the LRT for the first 10 days (n=4, 4 samples), post 10 days (n=4, 12 samples). Individual dots indicate independent samples, solid line indicates the mean and SEM. 2-way Anova was used to compare between groups. Pre-10 days ( Day 1 - Day 10), Post-10 days (Day 11- )

### Supplementary Figure 4

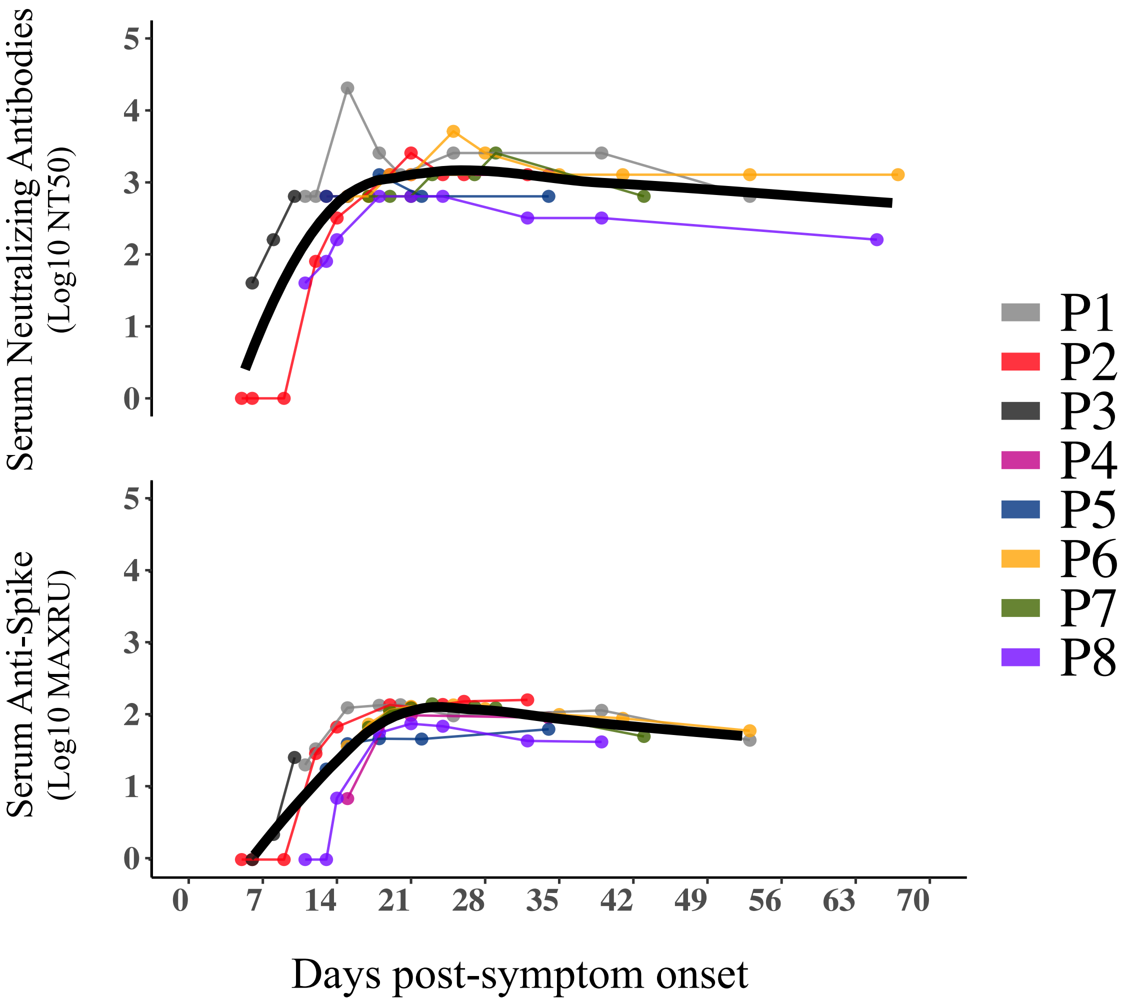


**Figure S4.** Serum Neutralizing Antibodies and Serum Anti-spike antibody levels in blood of severe COVID-19 patients. Data shown in logarithmic scale. Each patient is shown in different color. Black solid line shows the mean value.
