## Supplementary Tables for "Antiviral innate immunity is diminished in the upper respiratory tract of severe COVID-19 patients"

### Supplementary Table 1

| Patient Number | Age Range | Race | Charlson Score | Day post-symptom onset on admission | Day post-symptom onset on first URT sample | Max SOFA | Max Oxygen Support | Discharged or Died |
|---|---|---|---|---|---|---|---|---|
| 1 | 40-45 | White | 0 | 10 | 11 | 3 | HFNC | Discharged |
| 2 | 50-55 | White | 3 | 4 | 1 | 10 | MV | Discharged |
| 3 | 65-70 | Black/African American | 2 | 5 | 5 | 12 | MV | Died |
| 4 | 20-24 | White | 0 | 8 | 10 | 1 | HFNC | Discharged |
| 5 | 25-30 | Black/African American | 0 | 7 | 9 | 6 | MV | Discharged |
| 6 | 60-64 | Asian | 2 | 14 | 15 | 7 | MV | Discharged |
| 7 | 50-55 | White | 1 | 16 | 19 | 1 | Nasal canula | Discharged |
| 8 | 60-64 | Asian | 2 | 10 | 11 | 3 | HFNC | Discharged |

Table S1. Severe cohort description.

### Supplementary Table 2

| Patient | Remdesivir | Steroids | Tocilizumab |
|---|---|---|---|
| **1** | No | No | no |
| **2** | Yes | No | No |
| **3*** | Yes | Methylprednisolone | No |
| **4** | No | No | No |
| **5** | No | Methylprednisolone | Yes (disease day 11) |
| **6** | No | No | No |
| **7** | Yes | No | No |
| **8** | Yes | No | No |

*Patient died

Table S2. Severe Patients Treatments.

### Supplementary Table 3

| Patient | Age Range | Race | Day post-symptom onset on first URT sample |
|---|---|---|---|
| 1 | 20 - 25 | Asian | 5 |
| 2 | 30 - 35 | White | 1 |
| 3 | 20 - 25 | White | 2 |
| 4 | 36 - 40 | White | 5 |
| 5 | 20 - 25 | White | 7 |
| 6 | 55 - 60 | White | 16 |
| 7 | 26 - 30 | White | 6 |

Table S3. Mild Cohort Description.

### Supplementary Table 4

| Mild (first 10 days) | | | Mild (post-10 days) | | | Severe (first 10 days) | | | Severe (post-10 days) | | |
|---|---|---|---|---|---|---|---|---|---|---|---|
| Patient | Viral Load (copies/ml) | Days post-symptoms onset | Patient | Viral Load (copies/ml) | Days post-symptoms onset | Patient | Viral Load (copies/ml) | Days post-symptoms onset | Patient | Viral Load (copies/ml) | Days post-symptoms onset |
| **M1** | 2904472 | 5 | M1 | 389 | 13 | **S2** | 239.56 | 1 | S2 | 10483.64 | 11 |
| **M2** | 8708927 | 1 | | | | | 539.56 | 6 | | 19053.49 | 12 |
| | 5637 | 8 | | | | | 77402.67 | 7 | | 118.53 | 13 |
| **M3** | 4890344 | 2 | | | | | 1423569.87 | 8 | | 3596.57 | 14 |
| | 414 | 9 | | | | | 872984.06 | 9 | | 315.74 | 15 |
| **M4** | 8736696 | 5 | M4 | 1366 | 11 | | 1392050.16 | 10 | | 402.69 | 16 |
| **M5** | 89208 | 7 | M5 | 17790 | 14 | **S3** | 739.86 | 6 | S3 | 641864.78 | 11 |
| | | | | | | | 57.81 | 7 | | 42129.94 | 12 |
| | | | | | | | 2669499.66 | 8 | | | |
| **Average** | 3619385.429 | 5 | | 6515 | 13 | | 20231.51 | 9 | | | |
| | | | | | | | 10841.67 | 10 | | | |
| | | | | | | **S4** | 105735.39 | 10 | S4 | 3708253.04 | 11 |
| | | | | | | | | | | 159890.78 | 12 |
| | | | | | | | | | | 48123.28 | 14 |
| | | | | | | | | | | 1988.38 | 14 |
| | | | | | | | | | | 388.92 | 15 |
| | | | | | | | | | | 53.72 | 16 |
| | | | | | | **S5** | 3860244.05 | 9 | S5 | 281038.61 | 11 |
| | | | | | | | | | | 5863.65 | 12 |
| | | | | | | | | | | 4114.9 | 13 |
| | | | | | | | | | | 226967.07 | 14 |
| | | | | | | | | | | 1160.12 | 15 |
| | | | | | | | | | | 259.84 | 16 |
| | | | | | | **Average** | 802625.8331 | 8 | | 257803.3845 | 13 |

Table S4. Description of sample analyzed for pre and post day 10 comparisons

### Supplementary Table 5

| Marker* | CXCL10 | DDX60 | GBP1 | HERC5 | HERC6 | IFIT5 | OAS3 | OASL | RTP4 | SIGLEC1 | SOCS1 | SPATS2L | USP18 | LAMP3 (higher in LRT severe) |
|---|---|---|---|---|---|---|---|---|---|---|---|---|---|---|
| URT (mild) correlation with | IFN-I Score | IFN-I Score | IFN-I Score | *IFNG* | IFN-I Score | IFN-I Score | IFN-I Score | ns | IFN-I Score | NFkB Score, *IFNA2* | IFN-I Score | NFkB Score, *IFNA2* | NFkB Score, *IFNA2* | ns |
| URT (severe) correlation with | IFN-I Score | ns | ns | ns | IFN-I Score | ns | IFN-I Score | ns | ns | ns | ns | ns | ns | ns |
| LRT (severe) correlation with | IFN-I Score, NFKB score, *IFNG*, negative corr *IFNB1* | IFN-I Score, NFKB score, IFNG, negative corr *IFNB1* | IFN-I Score, NFKB score, IFNG, negative corr *IFNB1* | IFN-I Score, NFKB score, IFNG, negative corr *IFNB1* | IFN-I Score, NFKB score, IFNG, negative corr *IFNB1* | IFN-I Score, NFKB score, IFNG, negative corr *IFNB1* | IFN-I Score, NFKB score, IFNG, negative corr *IFNB1* | IFN-I Score, NFKB score, IFNG, negative corr *IFNB1* | IFN-I Score, NFKB score, IFNG, negative corr *IFNB1* | IFN-I Score, NFKB score, IFNG, negative corr *IFNB1* | IFN-I Score, NFKB score, IFNG, negative corr *IFNB1* | IFN-I Score, NFKB score, IFNG, negative corr *IFNB1* | IFN-I Score, NFKB score, *IFNG,* negative corr *IFNB1* | ns |
| Blood (severe) correlation with | NFKB score, IFNA2, IFNB1 | IFN-I Score | NFKB score, IFNB1 | IFN-I Score | IFN-I Score | IFN-I Score | IFN-I Score | IFN-I Score, NFKB score, IFNB1 | IFN-I Score, NFKB score, IFNB1 | IFN-I Score, IFNB1 | NFKB score, IFNA2, IFNB1 | IFN-I Score, NFKB score, IFNB1 | IFN-I Score, IFNB1 | IFN-I Score, NFKB score, IFNB1 |
| Function | Coordinates early adaptive immune responses through recruitment of CXCR3+ effector T and NK cells | Poorly defined | Poorly defined | Increases glycosylation | | Negatively regulated by disrupting TBK1-IKKe-IRF3 signaling | Interferon-inducible antiviral proteins | Promotes antiviral activity by enhancing the sensitivity of RIG-I activation | Potent IFN-inducible inhibitor of human pathogens in the Flaviviridae | Siglec-1 functions as attachment receptor by enhaning ACE2-mediated infection | Regulated IFNAR1- but not IFNAR2-specific signals, | Involved in restricting viral entry and replication | Enforces viral replication in DCs | LAMP3 is one of about 400 genes that are activated by the integrated stress response (ISR) downstream of ATF4 |
| Effect of low expression | Decreased might limit NK and T cell recruitment impairing recognition and clearance of SARS-CoV-2 infected cells | | | Reduced ISGylation increases viral stability | | Reduced expression leads to stabilized and higher TBK1-IFFe-IRF3 function | Less degradation of viral RNA, inhibition of virus replication | Lack of RIG-I activity may impair the early restraining of SARS-CoV-2 infection in human lung cells. | Less genome amplification and viral production | Reduced Siglec1 in macrophages/DC may reduce viral uptake and infection | Poor regulation of type I interferon signaling and less balances of its beneficial antiviral versus detrimental proinflammatory effects. | Reduced SPATS2L leads to unreduced high SARS-CoV-2 RNA levels and viral entry and replication are less restricted | High USP18 early in infection in monocytes and DC may induce viral replication necessary to mount an effective TH1 and CD8+ T cell response respectively | Elevated levels are suggestive of an increased integrated stress response |
| Reference | 1 | | | 2 | | 3 | **4** | **5,6** | **7** | **8** | **9** | **10** | 11,12 | **13** |

* All markers were higher in URT swabs from patients with mild disease compared to the URT swabs from severe patients except for LAMP3 which was lower

References:

| | |
|---|---|
| 1 | Liu M, Guo S, Hibbert JM, Jain V, Singh N, Wilson NO, Stiles JK. CXCL10/IP-10 in infectious diseases pathogenesis and potential therapeutic implications. Cytokine Growth Factor Rev. 2011 Jun;22(3):121-30. doi: 10.1016/j.cytogfr.2011.06.001. Epub 2011 Jul 29. PMID: 21802343; PMCID: PMC3203691. |
| 2 | Mathieu NA, Paparisto E, Barr SD, Spratt DE. HERC5 and the ISGylation Pathway: Critical Modulators of the Antiviral Immune Response. Viruses. 2021 Jun 9;13(6):1102. doi: 10.3390/v13061102. PMID: 34207696; PMCID: PMC8228270. |
| 3 | Zhang N, Shi H, Yan M, Liu G. IFIT5 Negatively Regulates the Type I IFN Pathway by Disrupting TBK1-IKKε-IRF3 Signalosome and Degrading IRF3 and IKKε. Journal of Immunology (Baltimore, Md. : 1950). 2021 May;206(9):2184-2197. DOI: 10.4049/jimmunol.2001033. PMID: 33858962. |
| 4 | Banday AR, Stanifer ML, Florez-Vargas O, Onabajo OO, Papenberg BW, Zahoor MA, Mirabello L, Ring TJ, Lee CH, Albert PS, Andreakos E, Arons E, Barsh G, Biesecker LG, Boyle DL, Brahier MS, Burnett-Hartman A, Carrington M, Chang E, Choe PG, Chisholm RL, Colli LM, Dalgard CL, Dude CM, Edberg J, Erdmann N, Feigelson HS, Fonseca BA, Firestein GS, Gehring AJ, Guo C, Ho M, Holland S, Hutchinson AA, Im H, Irby L, Ison MG, Joseph NT, Kim HB, Kreitman RJ, Korf BR, Lipkin SM, Mahgoub SM, Mohammed I, Paschoalini GL, Pacheco JA, Peluso MJ, Rader DJ, Redden DT, Ritchie MD, Rosenblum B, Ross ME, Anna HPS, Savage SA, Sharma S, Siouti E, Smith AK, Triantafyllia V, Vargas JM, Vargas JD, Verma A, Vij V, Wesemann DR, Yeager M, Yu X, Zhang Y, Boulant S, Chanock SJ, Feld JJ, Prokunina-Olsson L. Genetic regulation of OAS1 nonsense-mediated decay underlies association with COVID-19 hospitalization in patients of European and African ancestries. Nat Genet. 2022 Aug;54(8):1103-1116. doi: 10.1038/s41588-022-01113-z. Epub 2022 Jul 14. PMID: 35835913; PMCID: PMC9355882. |
| 5 | Zhu J, Ghosh A, Sarkar SN. OASL-a new player in controlling antiviral innate immunity. Curr Opin Virol. 2015 Jun;12:15-9. doi: 10.1016/j.coviro.2015.01.010. Epub 2015 Feb 9. PMID: 25676874; PMCID: PMC4470762. |
| 6 | Yamada, T., Sato, S., Sotoyama, Y. et al. RIG-I triggers a signaling-abortive anti-SARS-CoV-2 defense in human lung cells. Nat Immunol 22, 820–828 (2021). |
| 7 | Boys IN, Xu E, Mar KB, De La Cruz-Rivera PC, Eitson JL, Moon B, Schoggins JW. RTP4 Is a Potent IFN-Inducible Anti-flavivirus Effector Engaged in a Host-Virus Arms Race in Bats and Other Mammals. Cell Host Microbe. 2020 Nov 11;28(5):712-723.e9. doi: 10.1016/j.chom.2020.09.014. Epub 2020 Oct 27. PMID: 33113352; PMCID: PMC7666060. |
| 8 | Lempp, F.A., Soriaga, L.B., Montiel-Ruiz, M. et al. Lectins enhance SARS-CoV-2 infection and influence neutralizing antibodies. Nature 598, 342–347 (2021). https://doi.org/10.1038/s41586-021-03925-1 |
| 9 | Fenner JE, Starr R, Cornish AL, Zhang JG, Metcalf D, Schreiber RD, Sheehan K, Hilton DJ, Alexander WS, Hertzog PJ. Suppressor of cytokine signaling 1 regulates the immune response to infection by a unique inhibition of type I interferon activity. Nat Immunol. 2006 Jan;7(1):33-9. doi: 10.1038/ni1287. Epub 2005 Nov 27. PMID: 16311601. |
| 10 | Martin-Sancho L, Lewinski MK, Pache L, Stoneham CA, Yin X, Becker ME, Pratt D, Churas C, Rosenthal SB, Liu S, Weston S, De Jesus PD, O'Neill AM, Gounder AP, Nguyen C, Pu Y, Curry HM, Oom AL, Miorin L, Rodriguez-Frandsen A, Zheng F, Wu C, Xiong Y, Urbanowski M, Shaw ML, Chang MW, Benner C, Hope TJ, Frieman MB, García-Sastre A, Ideker T, Hultquist JF, Guatelli J, Chanda SK. Functional landscape of SARS-CoV-2 cellular restriction. Mol Cell. 2021 Jun 17;81(12):2656-2668.e8. doi: 10.1016/j.molcel.2021.04.008. Epub 2021 Apr 13. PMID: 33930332; PMCID: PMC8043580. |
| 11 | Honke N, Shaabani N, Zhang DE, Iliakis G, Xu HC, Häussinger D, Recher M, Löhning M, Lang PA, Lang KS. Usp18 driven enforced viral replication in dendritic cells contributes to break of immunological tolerance in autoimmune diabetes. PLoS Pathog. 2013 Oct;9(10):e1003650. doi: 10.1371/journal.ppat.1003650. Epub 2013 Oct 24. PMID: 24204252; PMCID: PMC3812017. |
| 12 | Ritchie, K., Hahn, C., Kim, K. et al. Role of ISG15 protease UBP43 (USP18) in innate immunity to viral infection. Nat Med 10, 1374–1378 (2004). https://doi.org/10.1038/nm1133 |
| 13 | Burton TD, Fedele AO, Xie J, Sandeman LY, Proud CG. The gene for the lysosomal protein LAMP3 is a direct target of the transcription factor ATF4. J Biol Chem. 2020 May 22;295(21):7418-7430. doi: 10.1074/jbc.RA119.011864. Epub 2020 Apr 20. PMID: 32312748; PMCID: PMC7247307. |

Table S5. Summary of the antiviral role of 13 genes with significant differences between mild and severe patients.
